# Multidimensional nutritional assessment in Crohn’s disease: cross-sectional comparison of active disease and remission

**DOI:** 10.64898/2026.06.11.26355434

**Authors:** Aditi Sarker, Chanchal Kumar Ghosh, Prodipta Chowdhury

## Abstract

Malnutrition is common in Crohn’s disease (CD), and its assessment requires multiple tools. Comprehensive evaluation of nutritional status in a population with CD, predominantly characterized by metabolic phenotype, was inadequately reported. This study evaluated the nutritional status of CD patients using anthropometric, clinical, and biochemical measures and compared patients with active disease with those in remission. This cross-sectional study included 127 adults with CD: 63 with active disease and 64 in remission. Disease activity was classified using the Crohn’s Disease Activity Index, the Simple Endoscopic Score for Crohn’s Disease, and magnetic resonance enterography. Nutritional assessment included body mass index (BMI), mid-upper arm circumference, calf circumference, triceps skinfold thickness, mid-arm muscle circumference, Mini Nutritional Assessment-Short Form (MNA-SF), and biochemical markers including hemoglobin, serum iron, folate, vitamin B12, albumin, and zinc. Malnutrition was defined using the Global Leadership Initiative on Malnutrition criteria. Overall, 47.2% of participants were malnourished. Malnutrition was significantly more frequent in active disease than in remission (81.0% vs. 14.1%, *P*<0.001). Patients with active CD had lower anthropometric indices, MNA-SF score, hemoglobin, serum iron, albumin, and zinc (all *P*<0.001), whereas folate and vitamin B12 did not differ significantly. BMI showed positive correlations with other anthropometric measures and MNA-SF score (r=0.854–0.914, all *P*<0.001), whereas correlations with biochemical parameters were weaker and disappeared after subgroup stratification. Overall, the findings indicate that malnutrition is highly prevalent in CD, particularly during active disease. Anthropometric measures and MNA-SF were strongly concordant, whereas biochemical markers were less consistent, supporting a multidimensional nutritional assessment approach in CD.

## INTRODUCTION

Crohn’s disease (CD) is a chronic, relapsing inflammatory bowel disease that can involve any part of the gastrointestinal tract. Malnutrition is a frequent but often underrecognized complication and, alongside inflammatory control, is an important determinant of clinical outcome [1,2].

In CD, nutritional impairment is multifactorial and may result from reduced intake, malabsorption, enteric nutrient loss, increased metabolic demand, and cytokine-driven catabolism. Consequently, patients may develop both protein-energy malnutrition and micronutrient deficiencies, including iron, folate, vitamin B12, and zinc, during active disease and even in remission, and these deficiencies may be associated with significant extraintestinal manifestations [3,4]. Studies have shown that patients with inflammatory bowel disease may continue to experience nutritional and functional deficits even after prolonged remission, despite apparently adequate macronutrient intake, emphasizing the need for routine nutritional assessment beyond symptomatic disease alone [3,4]. Assessment remains challenging because no single measure is sufficient in routine practice [5]. Anthropometric indices such as body mass index (BMI), mid-upper arm circumference (MUAC), calf circumference (CC), triceps skinfold thickness (TSF), and mid-arm muscle circumference (MAMC) are practical bedside tools for detecting macronutrient depletion. Hand-grip strength is also frequently used, although it is better considered a functional marker of muscle performance than a direct measure of nutritional status, and its cutoff values are still being refined [6]. In contrast, biochemical testing is needed to identify clinically relevant micronutrient deficiencies [7].

Several assessment tools are available, but disease-specific instruments, e.g., the IBD-NST, INTICO-2 CD questionnaire, and the Short Dietary Screener Questionnaire, may be less feasible in lower-resource settings. The Mini Nutritional Assessment-Short Form (MNA-SF), although originally developed and validated for older adults, may serve as a pragmatic screening step when followed by a disease-specific evaluation or a formal diagnostic framework [8]. In this context, the Global Leadership Initiative on Malnutrition (GLIM) has provided a standardized framework for diagnosing adult malnutrition. Since its introduction in 2019, the GLIM framework has provided a standardized approach to diagnosing adult malnutrition across a broad range of medical and surgical populations. It has also been applied to Chinese and European patients with CD; contemporary data from South Asian patients with CD remain limited [9].

Malnutrition in CD has been reported in approximately 20% to 80% of patients overall, with consistently higher rates during active disease than during remission [2,5]. However, reported prevalence varies considerably across study settings, disease severity, ethnicities, and methods used for nutritional assessment. Moreover, many previous studies have been conducted in hospitalized populations or high-income settings, and relatively few have evaluated nutritional status across both active and remission phases using a multidimensional approach that integrates objective disease activity assessment with anthropometric and biochemical evaluations [4,10,11].

This gap is particularly important in South Asia. South Asian populations often display a thin-fat, metabolically high-risk phenotype, with greater central adiposity and cardiometabolic risk at lower BMI, which may mask nutritional impairment when BMI is considered alone [12]. At the same time, available regional studies suggest altered body composition in CD, but multidimensional nutritional evaluation across active disease and remission is still inadequately reported in this region [10,13].

Accordingly, this study assessed the nutritional status of adults with Crohn’s disease using integrated anthropometric, biochemical, and clinical parameters and compared patients with active disease with those in remission. By addressing the regional evidence gap, it aims to inform nutritional evaluation and management of CD in resource-limited settings.

## METHODS

### 1. Ethics statement

We conducted this study in compliance with the principles of the Declaration of Helsinki. The study’s protocol was reviewed and approved by the Institutional Review Board (IRB) of Bangladesh Medical University, Shahbagh, Dhaka (approval no. 4764; 11 Jan 2024), and written informed consent was obtained from all participants. This study was reported in accordance with the Strengthening the Reporting of Observational Studies in Epidemiology (STROBE) statement for cross-sectional studies; the completed checklist is provided as S1 Checklist.

### 2. Study Design and Participants

This cross-sectional observational study was conducted in the inpatient and outpatient Departments of Gastroenterology of a tertiary care hospital in Dhaka, Bangladesh. Adults aged 18 years or older with previously diagnosed or newly diagnosed Crohn’s disease were recruited through consecutive sampling from February 2024 to January 2025, following IRB approval. Sex was recorded as a biological variable and was self-reported by the participants at enrollment. Diagnosis was established from compatible clinical, endoscopic, histologic, and imaging findings. Patients were excluded if they were pregnant or had another condition that could independently affect nutritional status, including chronic liver disease, nephrotic syndrome, chronic pancreatitis, diabetes mellitus, tuberculosis, thyroid disorders, advanced cardiopulmonary disease, chronic kidney disease, or malignancy. The remission group served as the comparison group.

### 3. Disease Activity Assessment

Disease activity was classified using three complementary modalities: the Crohn’s Disease Activity Index (CDAI) for clinical activity, the Simple Endoscopic Score for Crohn’s Disease (SES-CD) for endoscopic activity, and magnetic resonance enterography (MRE) using a qualitative MR activity index for imaging activity [14–17]. Patients were categorized as having active disease if activity was present on any one of these assessments. Remission required the absence of activity across all three modalities.

### 4. Nutritional Assessment

Nutritional status was evaluated using anthropometry, the Mini Nutritional Assessment-Short Form (MNA-SF), and biochemical measurements. Height was measured to the nearest 0.1 cm using a wall-mounted stadiometer, and body weight was measured with minimal clothing using a calibrated electronic scale. Body mass index (BMI) was calculated as weight in kilograms divided by height in meters squared. Mid-upper arm circumference (MUAC) and calf circumference (CC) were measured with a non-stretch tape to the nearest 0.1 cm. Triceps skinfold thickness (TSF) was measured at the midpoint between the acromion and olecranon using skinfold calipers, and mid-arm muscle circumference (MAMC) was derived as MUAC - (3.14 x TSF). Measurements were obtained on the non-dominant arm using a standardized technique. A short dietary history was incorporated into the MNA-SF questionnaire.

### 5. Definition of Malnutrition

Malnutrition was defined using the Global Leadership Initiative on Malnutrition (GLIM) framework [18]. In accordance with the study protocol, patients were considered malnourished when at least one phenotypic criterion was present - low BMI according to Asian cutoffs, documented weight loss, or reduced muscle mass inferred from standard anthropometric measures - and the etiologic GLIM domain was fulfilled by Crohn’s disease-related inflammatory and gastrointestinal burden. Because reliable prior weight history was not available in all participants, severity staging could not be applied consistently; therefore, GLIM was operationalized dichotomously as malnutrition present or absent. Serum albumin and micronutrient concentrations were not used as diagnostic criteria for malnutrition and were analyzed separately as biochemical variables.

### 6. Biochemical and Imaging Assessment

Hemoglobin was measured using a Sysmex XN2000 six-part differential automated hematology analyzer in the institutional hematology laboratory. Serum zinc was analyzed using an Indiko Plus analyzer, and serum vitamin B12 and folate were measured using the Alinity ci analyzer in the institutional biochemistry laboratory. The remaining biochemical assays, including serum iron and albumin, were performed using the Atellica chemistry system (Siemens). Colonoscopy was performed in the institutional gastroenterology unit using Olympus CV 170 equipment, and MRE was performed in the institutional radiology department using the Magnetom Sempra system (Siemens).

### 7. Sample Size

The required sample size was estimated based on the previously reported between-group difference in BMI among patients with Crohn’s disease [19]. The calculation yielded a minimum of 63 patients in each group. A total of 127 patients were ultimately enrolled, including 63 with active disease and 64 in remission. The sample size was calculated using the following formula [20].

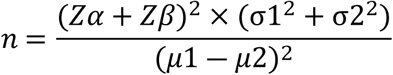

Here, Zα = The critical value of the Normal distribution at α/2 = 1.96 (For a confidence level of 95%, α is 0.05); Zβ = The critical value of the Normal distribution at β = 1.64 (For a power of 95%, β is 0.05); *μ*1 = The mean of BMI in remission group = 21.6; *μ*2 = The mean of BMI in active group = 18.8; σ1 = standard deviation of BMI in the remission group = 5; σ2 = standard deviation of BMI in the active group = 3.6. Using this formula, the minimum required sample in each group was 63.

### 8. Statistical Analysis

Data were processed using SPSS version 25.0 (IBM Corporation). Normality of continuous variables was assessed using the Shapiro-Wilk test and visual inspection of histograms and Q-Q plots, and statistical tests were selected accordingly. Analyses were performed using available complete data; missing values were not imputed, extreme values were checked for data-entry errors and retained when clinically plausible, and no formal correction for multiple comparisons was applied because the analyses were exploratory. Categorical variables were summarized as frequencies and percentages, and continuous variables as mean ± SD/SE or median with interquartile range, as appropriate. Comparison between the two groups (Active vs. remission) was performed using the Chi-square test for categorical data. Continuous variables were compared between groups using the unpaired Student’s t-test for normally distributed values and the Mann-Whitney U test for skewed values as applicable. Correlations between BMI and anthropometric, MNA-SF, and biochemical variables were examined using Pearson’s or Spearman’s correlation coefficients, as appropriate. All tests were two-sided, and *P*<0.05 was considered statistically significant.

## RESULTS

### 1. Baseline Characteristics of Study Participants

Table 1 compares the baseline characteristics of patients with CD during the active and remission phases. Sex distribution and the presence of extraintestinal manifestations were different between groups. 65.1% of males (n=41) were in the active group vs. 46.9% (n=30) in the remission group, whereas females in the active group were 34.9% (n=22) vs. 53.1% (n=34) in the remission group (*P*=0.039). Extraintestinal manifestations were more frequent in the active group than in the remission group [63.5% (n=40) vs. 26.6% (n=17), *P*=0.001]. Age, occupation, residence, income, smoking status, disease duration, disease location, disease behavior, presence of perianal disease, and frequency of relapse were similar between groups.

**Table 1.**
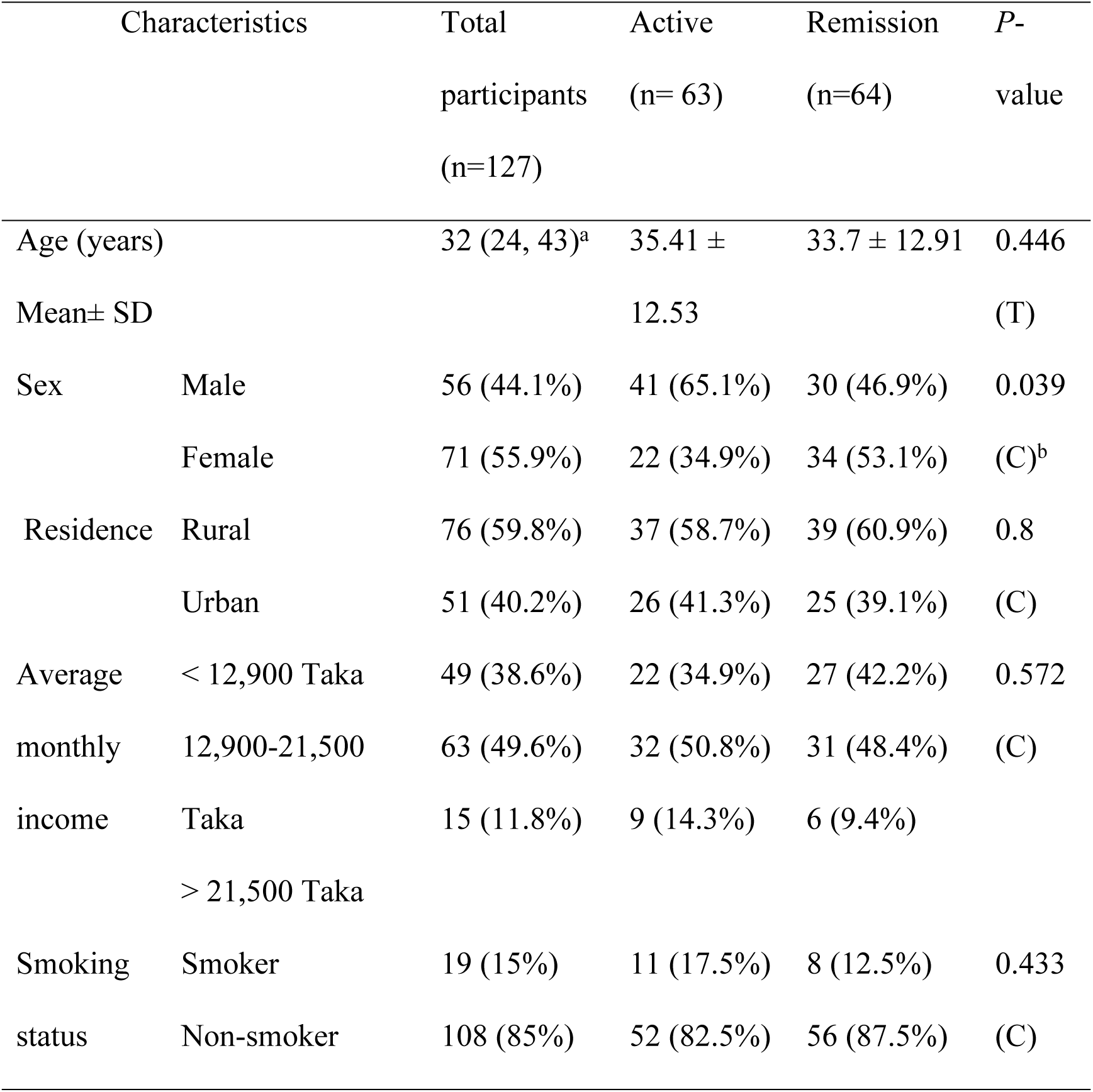
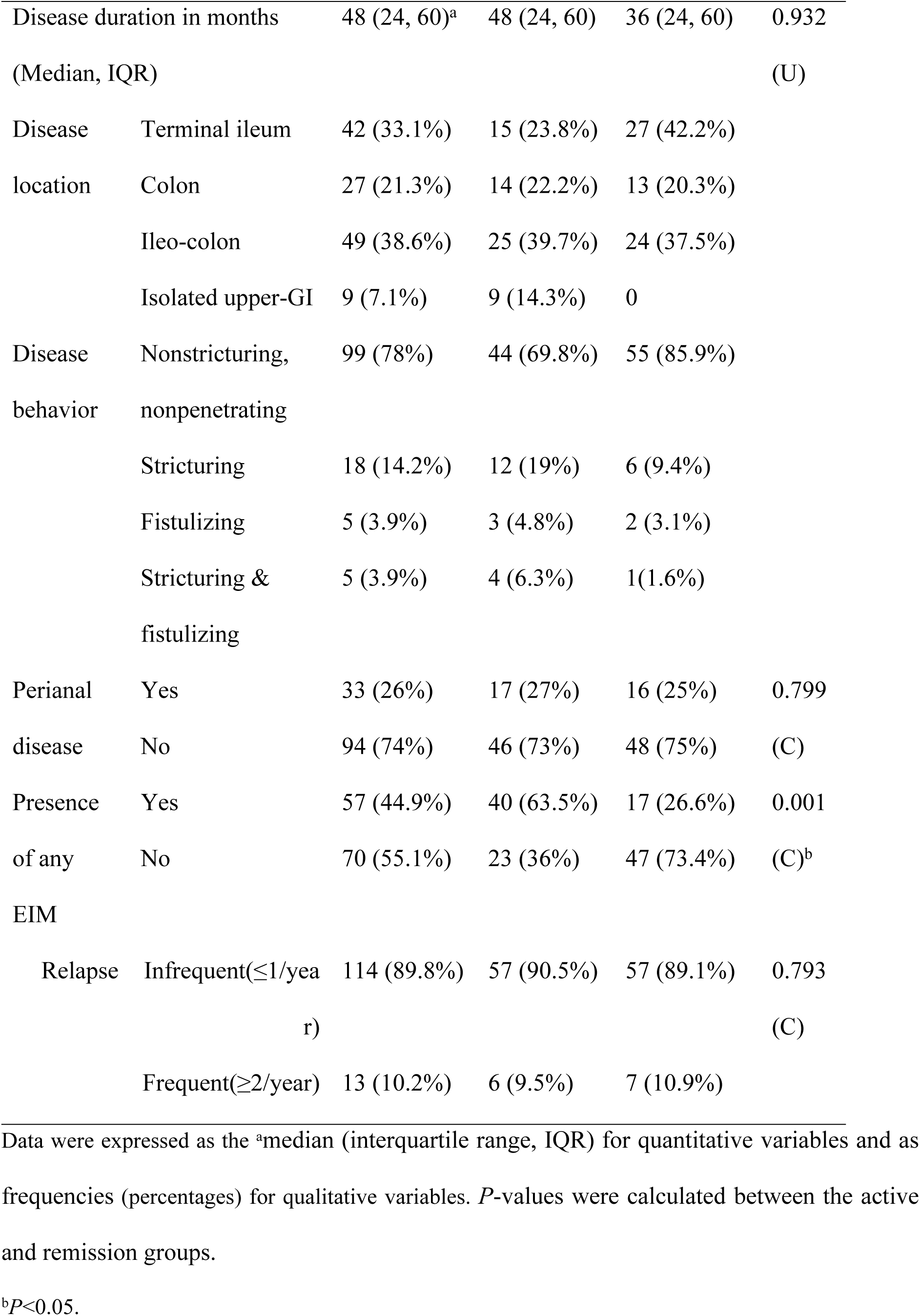
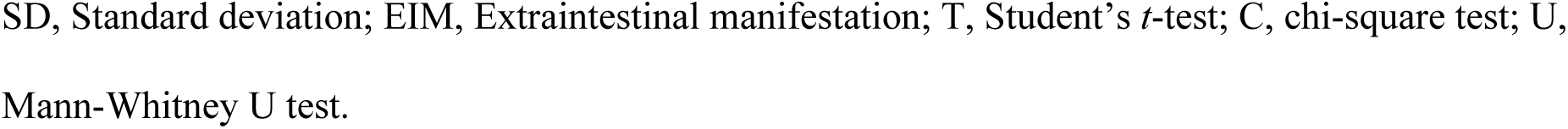
Baseline Characteristics of Study Participants (N=127)

| Characteristics |  | Total<br>participants<br>(n=127) | Active<br>(n= 63) | Remission<br>(n=64) | *P*-<br>value |
| --- | --- | --- | --- | --- | --- |
| Age (years) |  | 32 (24, 43)<sup>a</sup> | 35.41 ± | 33.7 ± 12.91 | 0.446 |
|  | Mean± SD |  | 12.53 |  | (T) |
| Sex | Male | 56 (44.1%) | 41 (65.1%) | 30 (46.9%) | 0.039 |
|  | Female | 71 (55.9%) | 22 (34.9%) | 34 (53.1%) | (C)<sup>b</sup> |
| Residence | Rural | 76 (59.8%) | 37 (58.7%) | 39 (60.9%) | 0.8 |
|  | Urban | 51 (40.2%) | 26 (41.3%) | 25 (39.1%) | (C) |
| Average<br>monthly<br>income | < 12,900 Taka | 49 (38.6%) | 22 (34.9%) | 27 (42.2%) | 0.572 |
|  | 12,900-21,500 | 63 (49.6%) | 32 (50.8%) | 31 (48.4%) | (C) |
|  | Taka<br>> 21,500 Taka | 15 (11.8%) | 9 (14.3%) | 6 (9.4%) |  |
| Smoking<br>status | Smoker | 19 (15%) | 11 (17.5%) | 8 (12.5%) | 0.433 |
|  | Non-smoker | 108 (85%) | 52 (82.5%) | 56 (87.5%) | (C) |
| Disease duration in months |  | 48 (24, 60)<sup>a</sup> | 48 (24, 60) | 36 (24, 60) | 0.932 |
| (Median, IQR) |  |  |  |  | (U) |
| Disease location | Terminal ileum | 42 (33.1%) | 15 (23.8%) | 27 (42.2%) |  |
|  | Colon | 27 (21.3%) | 14 (22.2%) | 13 (20.3%) |  |
|  | Ileo-colon | 49 (38.6%) | 25 (39.7%) | 24 (37.5%) |  |
|  | Isolated upper-GI | 9 (7.1%) | 9 (14.3%) | 0 |  |
| Disease behavior | Nonstricturing, nonpenetrating | 99 (78%) | 44 (69.8%) | 55 (85.9%) |  |
|  | Stricturing | 18 (14.2%) | 12 (19%) | 6 (9.4%) |  |
|  | Fistulizing | 5 (3.9%) | 3 (4.8%) | 2 (3.1%) |  |
|  | Stricturing & fistulizing | 5 (3.9%) | 4 (6.3%) | 1(1.6%) |  |
| Perianal disease | Yes | 33 (26%) | 17 (27%) | 16 (25%) | 0.799 |
|  | No | 94 (74%) | 46 (73%) | 48 (75%) | (C) |
| Presence of any EIM | Yes | 57 (44.9%) | 40 (63.5%) | 17 (26.6%) | 0.001 |
|  | No | 70 (55.1%) | 23 (36%) | 47 (73.4%) | (C)<sup>b</sup> |
| Relapse | Infrequent(≤1/year) | 114 (89.8%) | 57 (90.5%) | 57 (89.1%) | 0.793 |
|  |  |  |  |  | (C) |
|  | Frequent(≥2/year) | 13 (10.2%) | 6 (9.5%) | 7 (10.9%) |  |

Data were expressed as the <sup>a</sup>median (interquartile range, IQR) for quantitative variables and as frequencies (percentages) for qualitative variables. *P*-values were calculated between the active and remission groups.
<sup>b</sup>*P*<0.05.
SD, Standard deviation; EIM, Extraintestinal manifestation; T, Student's *t*-test; C, chi-square test; U, Mann-Whitney U test.

### 2. Anthropometric Assessment

Table 2 shows anthropometric parameters (BMI, MUAC, CC, TSF, MAMC) and the MNA-SF score for CD patients, and compares them between the active and remission groups. Patients with active disease had significantly lower body mass index, mid-upper arm circumference, calf circumference, triceps skinfold thickness, mid-arm muscle circumference, and Mini Nutritional Assessment-Short Form score than those in remission (all *P*<0.001).

**Table 2.**
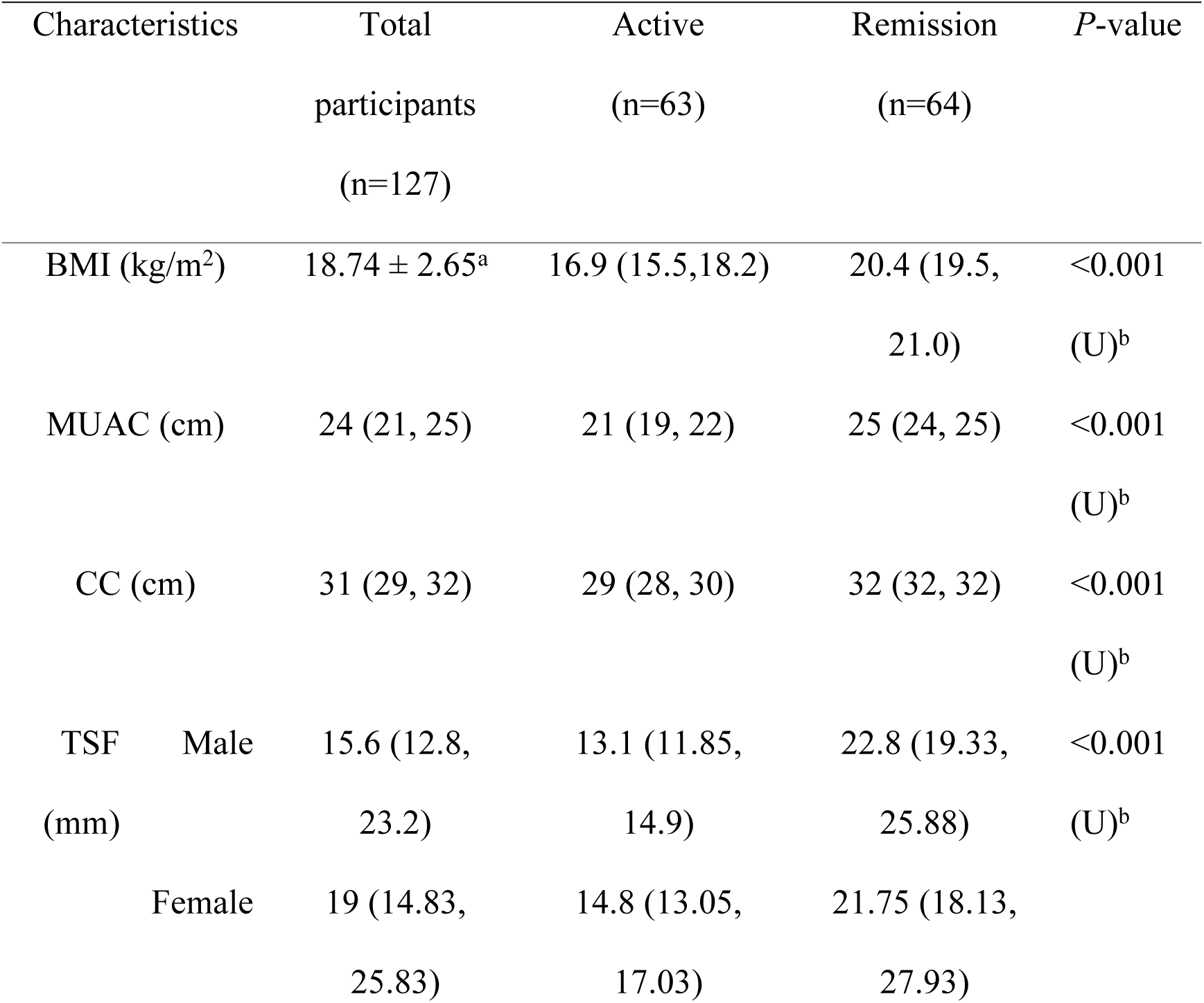
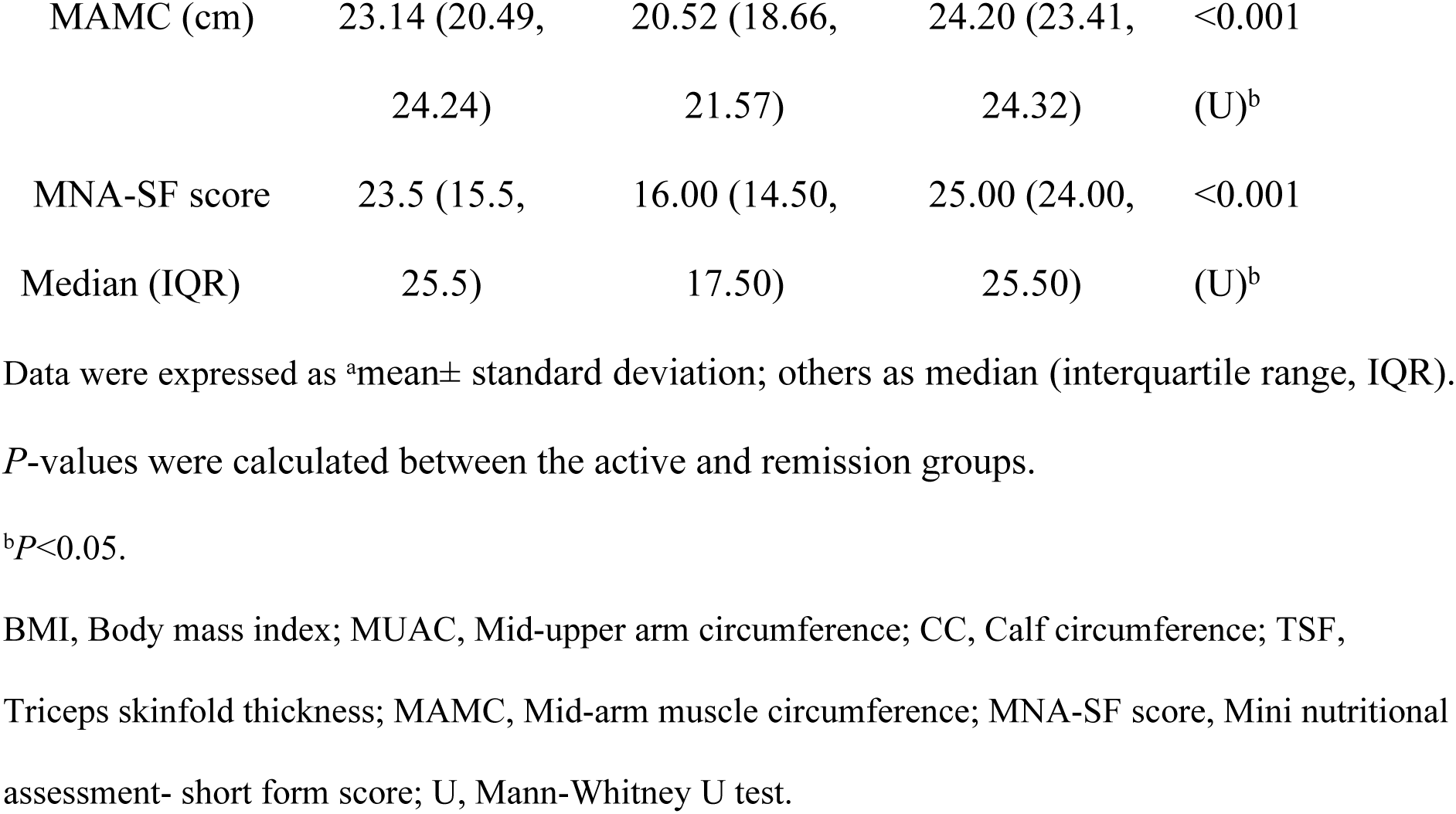
Anthropometric Assessment of Study Participants (N=127)

| Characteristics |  | Total<br>participants<br>(n=127) | Active<br>(n=63) | Remission<br>(n=64) | *P*-value |
| --- | --- | --- | --- | --- | --- |
| BMI (kg/m<sup>2</sup>) |  | 18.74 ± 2.65<sup>a</sup> | 16.9 (15.5, 18.2) | 20.4 (19.5, 21.0) | <0.001<br>(U)<sup>b</sup> |
| MUAC (cm) |  | 24 (21, 25) | 21 (19, 22) | 25 (24, 25) | <0.001<br>(U)<sup>b</sup> |
| CC (cm) |  | 31 (29, 32) | 29 (28, 30) | 32 (32, 32) | <0.001<br>(U)<sup>b</sup> |
| TSF (mm) | Male | 15.6 (12.8, 23.2) | 13.1 (11.85, 14.9) | 22.8 (19.33, 25.88) | <0.001<br>(U)<sup>b</sup> |
|  | Female | 19 (14.83, 25.83) | 14.8 (13.05, 17.03) | 21.75 (18.13, 27.93) |  |

|  |  |  |  |  |
| --- | --- | --- | --- | --- |
| MAMC (cm) | 23.14 (20.49, 24.24) | 20.52 (18.66, 21.57) | 24.20 (23.41, 24.32) | <0.001 (U)<sup>b</sup> |
| MNA-SF score | 23.5 (15.5, 25.5) | 16.00 (14.50, 17.50) | 25.00 (24.00, 25.50) | <0.001 (U)<sup>b</sup> |

Data were expressed as <sup>a</sup>mean± standard deviation; others as median (interquartile range, IQR).
*P*-values were calculated between the active and remission groups.
<sup>b</sup>*P*<0.05.
BMI, Body mass index; MUAC, Mid-upper arm circumference; CC, Calf circumference; TSF,
Triceps skinfold thickness; MAMC, Mid-arm muscle circumference; MNA-SF score, Mini nutritional
assessment- short form score; U, Mann-Whitney U test.

### 3. Nutritional Classification by GLIM criteria

According to the GLIM criteria, 81.0% of patients with active disease were classified as malnourished, compared with 14.1% of those in remission (Table 3; *P*<0.001).

**Table 3.** Classification of Study Participants (N=127) According to GLIM Criteria.

| Characteristics | Active<br>(n=63) | Remission<br>(n=64) | *P*-value |
| --- | --- | --- | --- |
| Normal nutritional status | 12 (19%) | 55 (85.9%) | <0.001 |
| Malnutrition | 51 (81%) | 9 (14.1%) | (C)<sup>a</sup> |

Data were expressed as frequencies (percentages, over row total)
<sup>a</sup>*P*<0.05.
C, Chi-square test

### 4. Biochemical Parameters

Table 4 summarizes biochemical parameters. Hemoglobin, serum iron, albumin, and zinc concentrations were significantly lower in active disease than in remission (all *P*<0.001, except hemoglobin in females, *P*=0.001). Folate and vitamin B12 did not differ significantly between groups. Table 5 presents all biochemical parameters, categorized as normal or low according to the cutoff, revealing substantial differences between nutritional groups classified by the GLIM criteria.

**Table 4.**
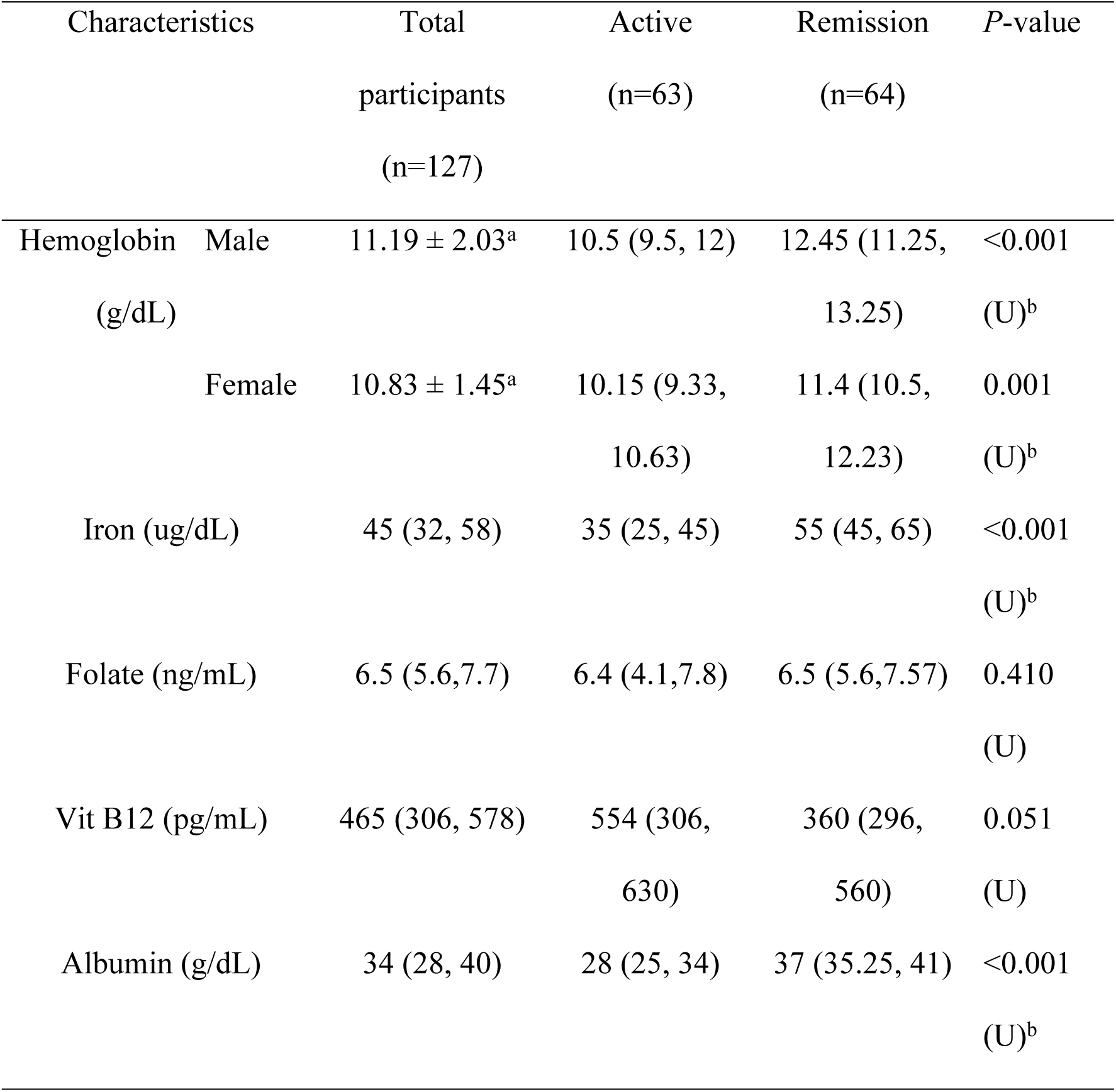
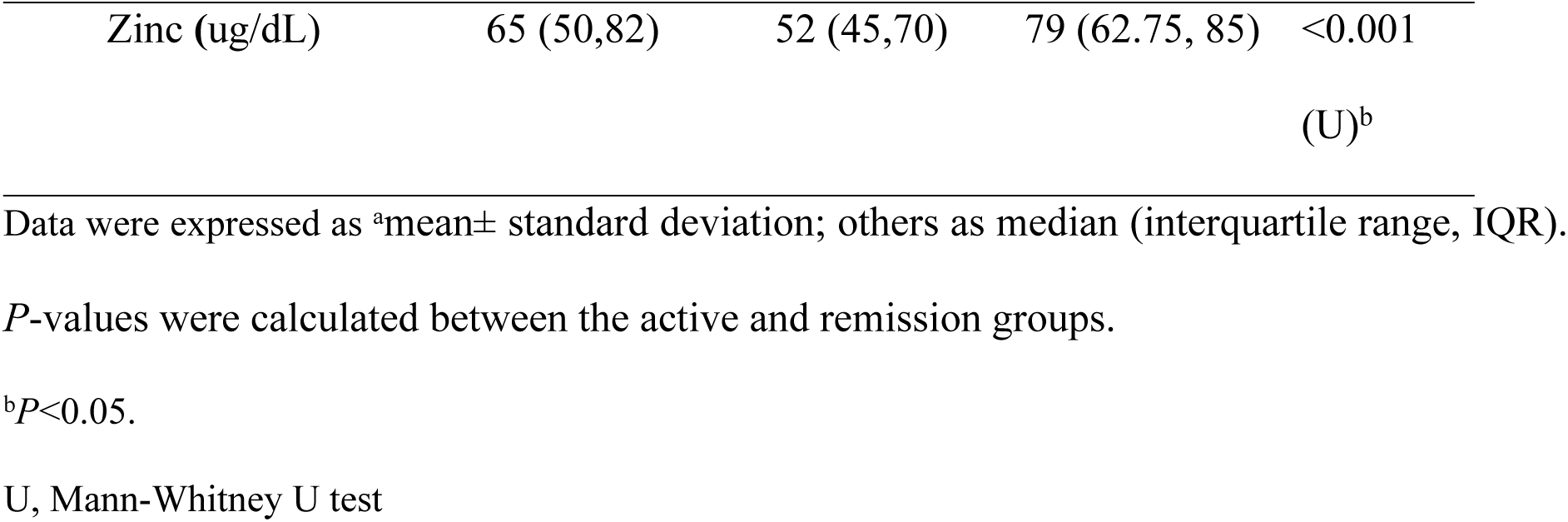
Biochemical Parameters of Study Participants (N=127)

| Characteristics |  | Total<br>participants<br>(n=127) | Active<br>(n=63) | Remission<br>(n=64) | P-value |
| --- | --- | --- | --- | --- | --- |
| Hemoglobin<br>(g/dL) | Male | 11.19 ± 2.03<sup>a</sup> | 10.5 (9.5, 12) | 12.45 (11.25, 13.25) | <0.001<br>(U)<sup>b</sup> |
|  | Female | 10.83 ± 1.45<sup>a</sup> | 10.15 (9.33, 10.63) | 11.4 (10.5, 12.23) | 0.001<br>(U)<sup>b</sup> |
| Iron (ug/dL) |  | 45 (32, 58) | 35 (25, 45) | 55 (45, 65) | <0.001<br>(U)<sup>b</sup> |
| Folate (ng/mL) |  | 6.5 (5.6, 7.7) | 6.4 (4.1, 7.8) | 6.5 (5.6, 7.57) | 0.410<br>(U) |
| Vit B12 (pg/mL) |  | 465 (306, 578) | 554 (306, 630) | 360 (296, 560) | 0.051<br>(U) |
| Albumin (g/dL) |  | 34 (28, 40) | 28 (25, 34) | 37 (35.25, 41) | <0.001<br>(U)<sup>b</sup> |

|  |  |  |  |  |
| --- | --- | --- | --- | --- |
| Zinc (ug/dL) | 65 (50,82) | 52 (45,70) | 79 (62.75, 85) | <0.001 |
|  |  |  |  | (U)<sup>b</sup> |

Data were expressed as <sup>a</sup>mean± standard deviation; others as median (interquartile range, IQR).
*P*-values were calculated between the active and remission groups.
<sup>b</sup>*P*<0.05.
U, Mann-Whitney U test

**Table 5.**
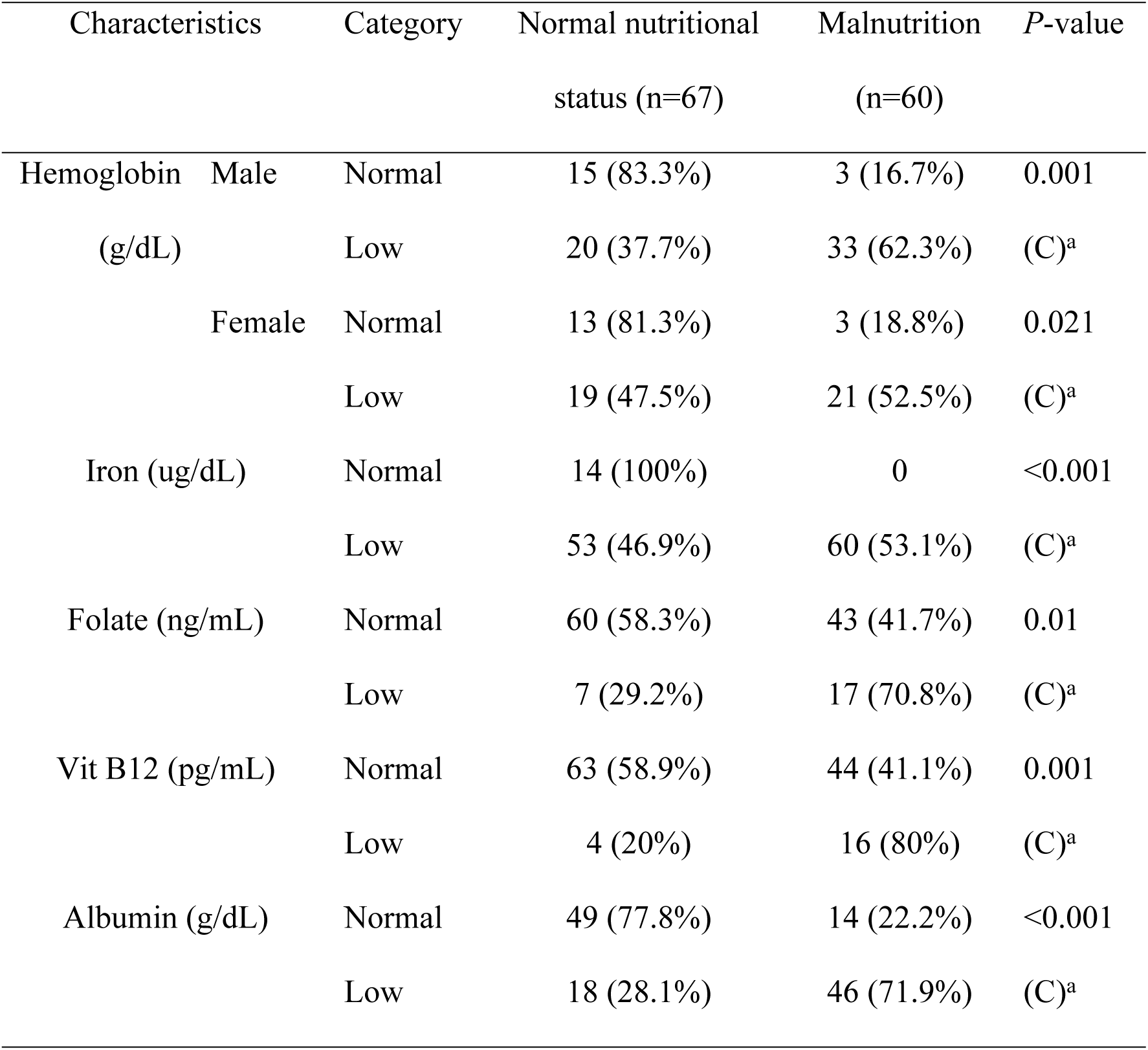
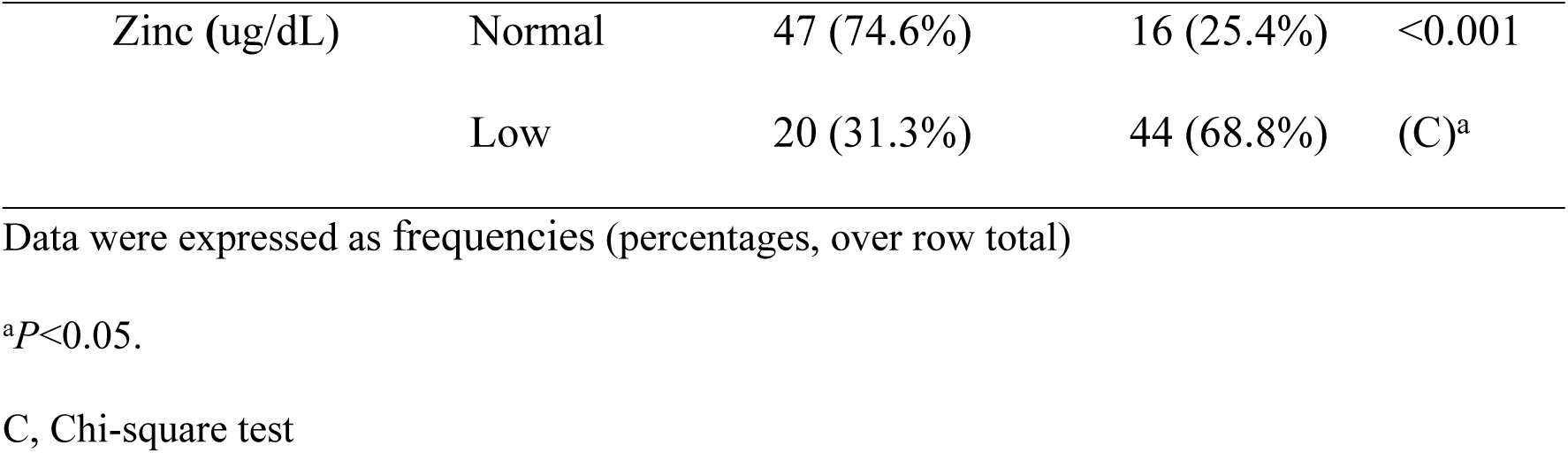
Category of Study Participants (N=127) By Different Biochemical Parameters and Comparison Between Nutritional Groups Classified According to GLIM Criteria.

| Characteristics |  | Category | Normal nutritional status (n=67) | Malnutrition (n=60) | *P*-value |
| --- | --- | --- | --- | --- | --- |
| Hemoglobin (g/dL) | Male | Normal | 15 (83.3%) | 3 (16.7%) | 0.001 |
|  |  | Low | 20 (37.7%) | 33 (62.3%) | (C)<sup>a</sup> |
|  | Female | Normal | 13 (81.3%) | 3 (18.8%) | 0.021 |
|  |  | Low | 19 (47.5%) | 21 (52.5%) | (C)<sup>a</sup> |
| Iron (ug/dL) |  | Normal | 14 (100%) | 0 | <0.001 |
|  |  | Low | 53 (46.9%) | 60 (53.1%) | (C)<sup>a</sup> |
| Folate (ng/mL) |  | Normal | 60 (58.3%) | 43 (41.7%) | 0.01 |
|  |  | Low | 7 (29.2%) | 17 (70.8%) | (C)<sup>a</sup> |
| Vit B12 (pg/mL) |  | Normal | 63 (58.9%) | 44 (41.1%) | 0.001 |
|  |  | Low | 4 (20%) | 16 (80%) | (C)<sup>a</sup> |
| Albumin (g/dL) |  | Normal | 49 (77.8%) | 14 (22.2%) | <0.001 |
|  |  | Low | 18 (28.1%) | 46 (71.9%) | (C)<sup>a</sup> |

|  |  |  |  |  |
| --- | --- | --- | --- | --- |
| Zinc (ug/dL) | Normal | 47 (74.6%) | 16 (25.4%) | <0.001 |
|  | Low | 20 (31.3%) | 44 (68.8%) | (C)<sup>a</sup> |

Data were expressed as frequencies (percentages, over row total)
<sup>a</sup> $P < 0.05$ .
C, Chi-square test

### 5. Correlation of BMI With Other Anthropometric Parameters and The MNA-SF Score

Table 6 shows the correlations between BMI and other anthropometric parameters and the MNA-SF score among study participants (n=127). BMI shows a very strong correlation with other anthropometric parameters, such as MUAC (r=0.88), CC (r=0.854), TSF (r=0.914), and MAMC (r=0.868), as well as with the MNA-SF score (r=0.876) (all *P*<0.001).

**Table 6.**
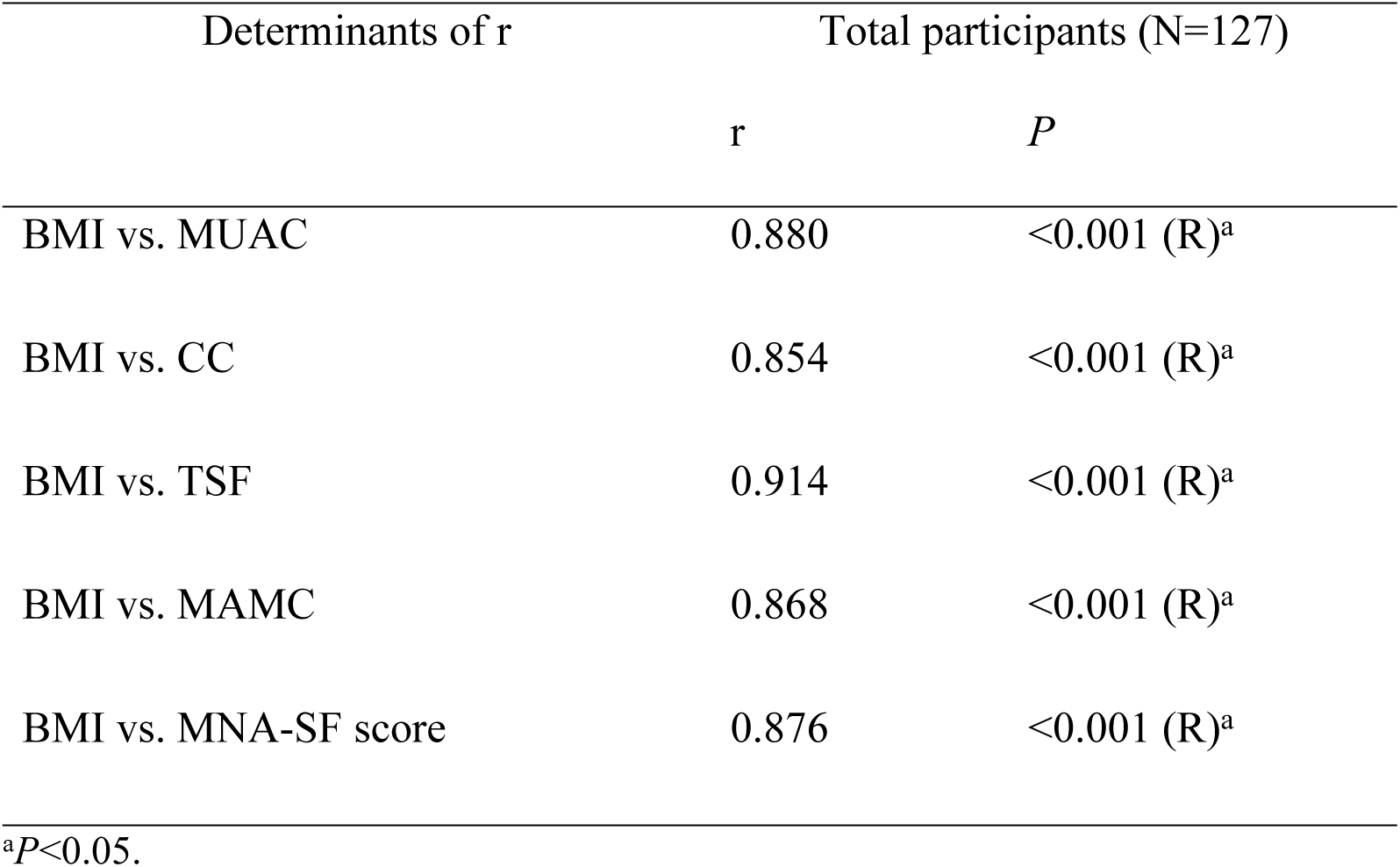
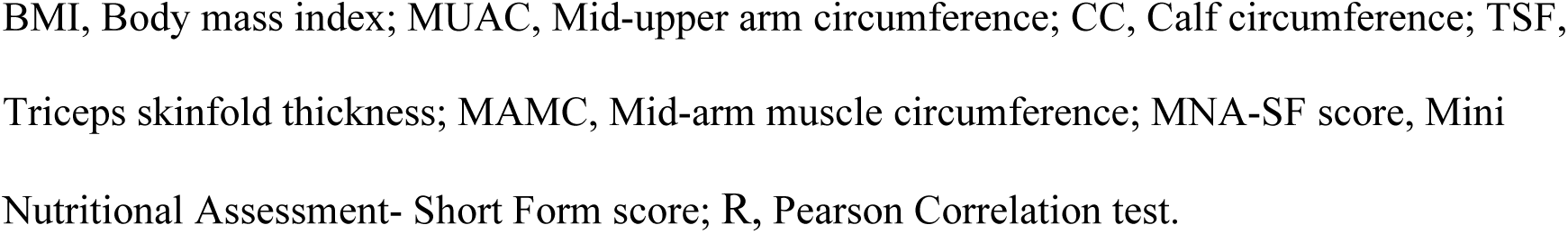
Correlation of BMI With Other Anthropometric Parameters and MNA-SF Score of Study Participants (N=127)

| Determinants of r | Total participants (N=127) |  |
| --- | --- | --- |
|  | r | P |
| BMI vs. MUAC | 0.880 | <0.001 (R)<sup>a</sup> |
| BMI vs. CC | 0.854 | <0.001 (R)<sup>a</sup> |
| BMI vs. TSF | 0.914 | <0.001 (R)<sup>a</sup> |
| BMI vs. MAMC | 0.868 | <0.001 (R)<sup>a</sup> |
| BMI vs. MNA-SF score | 0.876 | <0.001 (R)<sup>a</sup> |

<sup>a</sup> $P < 0.05$ .
BMI, Body mass index; MUAC, Mid-upper arm circumference; CC, Calf circumference; TSF, Triceps skinfold thickness; MAMC, Mid-arm muscle circumference; MNA-SF score, Mini Nutritional Assessment- Short Form score; R, Pearson Correlation test.

### 6. Correlation of BMI With Biochemical Parameters

Overall, body mass index was significantly correlated with hemoglobin, serum iron, albumin, and zinc, although the associations were weak to moderate in magnitude (Table 7). After stratification by disease activity and nutritional group, most correlations were no longer significant. The only significant subgroup correlations were with hemoglobin in the remission group (r_s_=0.267, *P*=0.033) and in the malnutrition group (r_s_=-0.267, *P*=0.039).

**Table 7.**
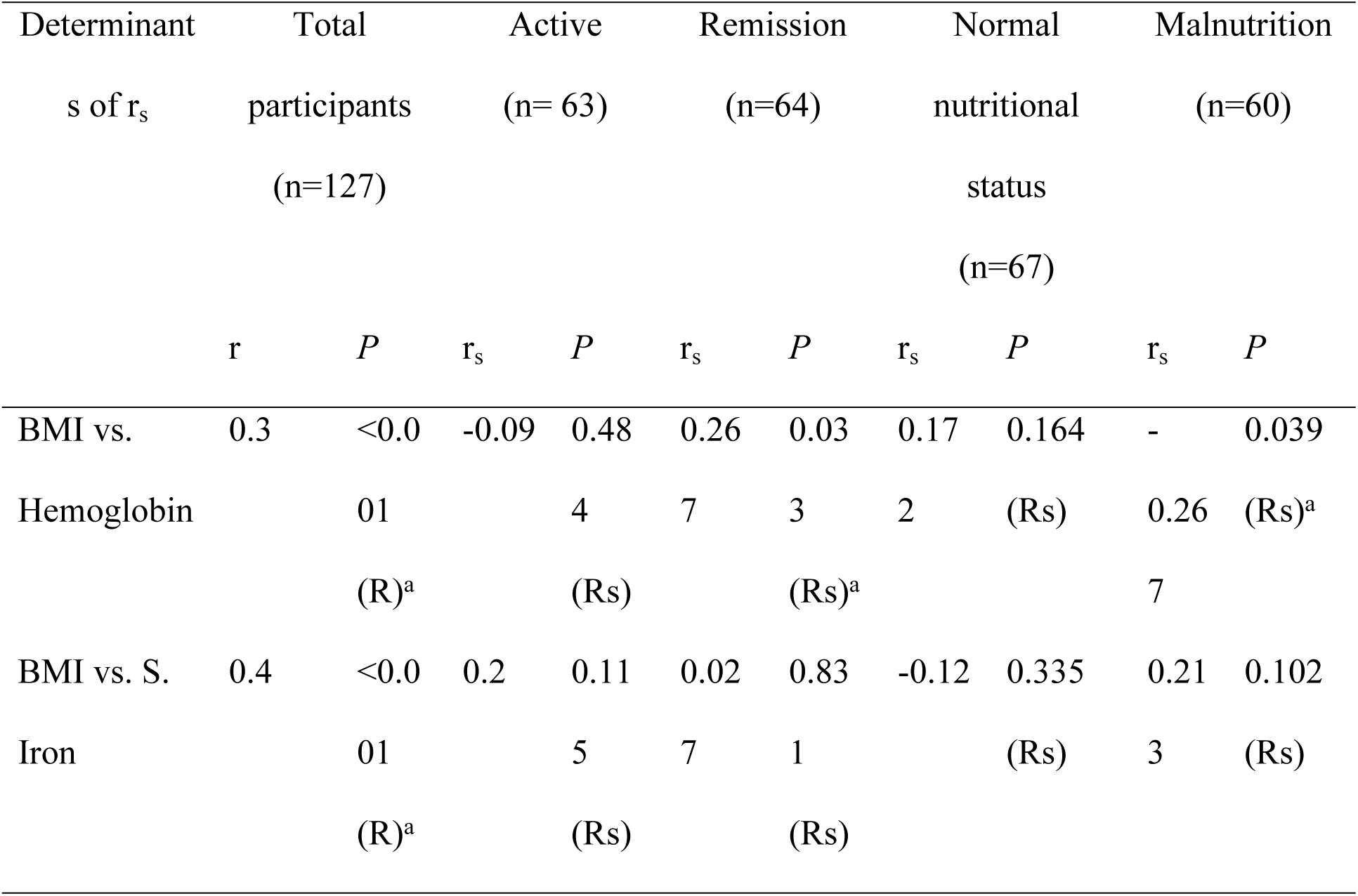
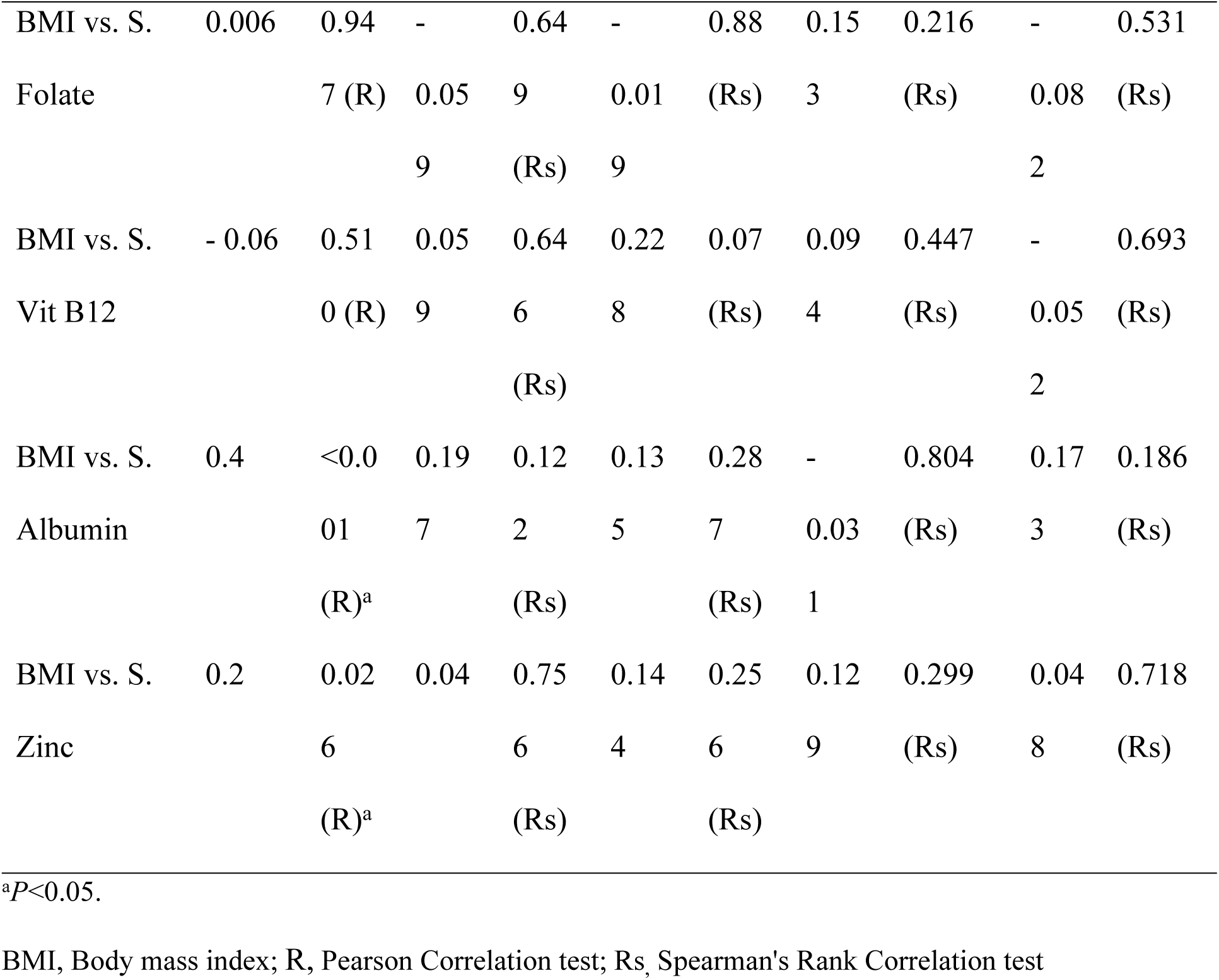
Correlation of BMI With Biochemical Parameters of Study Participants (N=127)

## DISCUSSION

This cross-sectional study evaluated the nutritional status of adults with Crohn’s disease during active and remission phases using a multimodal approach that included GLIM-based classification, anthropometry, questionnaire-based screening, and selected biochemical markers. Nearly half of the participants met the criteria for malnutrition, and the burden was predominantly among patients with active disease. In parallel, active disease was associated with a poorer anthropometric profile and lower concentrations of hemoglobin, serum iron, albumin, and zinc. BMI tracked closely with other anthropometric measures and the MNA-SF score, whereas its associations with biochemical indices were weaker and mostly disappeared after subgroup stratification. Taken together, these findings suggest that nutritional impairment in CD is strongly linked to the disease itself, persists throughout both the active and remission phases, and is best characterized by a multidimensional assessment strategy rather than any single nutritional marker.

The observed frequency of malnutrition is broadly consistent with previous literature reporting malnutrition in approximately 20% to 80% of patients with CD, with higher rates during active disease than during remission [2,5]. The marked difference between the active and remission phases in the present study is also consistent with reports from India and Brazil, where poorer nutritional status has been described among patients with active disease [10,19]. This pattern is biologically plausible because active inflammation may simultaneously reduce intake, impair absorption, increase enteric nutrient loss, and intensify cytokine-mediated catabolism [3,4]. At the same time, the presence of malnutrition in a substantial proportion of patients in remission suggests that nutritional compromise may persist beyond overt inflammatory activity, supporting the need for surveillance even during the disease’s remission phase [4,21]. The significantly higher frequency of extraintestinal manifestations in the active group is also noteworthy and is consistent with reports that several extraintestinal features parallel intestinal inflammatory activity [22]. Although causal pathways cannot be established in a cross-sectional design, the co-occurrence of active disease, poorer nutritional status, and a greater burden of systemic manifestations underscores the clinical value of integrating nutritional assessment into the overall evaluation of disease.

In this study, the anthropometric parameters (e.g., BMI, MUAC, CC, TSF, and MAMC) were significantly poorer in active disease, and BMI showed very strong correlations with the other anthropometric variables and the MNA-SF score. These findings support the utility of simple bedside assessments, such as BMI, in routine CD care, particularly in resource-limited settings where disease-specific dietary tools or advanced body composition techniques may not be readily available. Despite the strong correlations observed in this group, the study also showed that BMI alone is not sufficient. BMI does not correlate with the full spectrum of micronutrient insufficiency. It is therefore better regarded as an entry-point measure within a broader nutritional assessment framework rather than as a standalone indicator of nutritional status [5,8]. In addition, a substantial proportion of CD patients categorized as normal by the GLIM criteria exhibited biochemical deficiencies, emphasizing that GLIM alone is insufficient as a marker of malnutrition in CD.

The biochemical profile reflected a greater burden of nutritional and inflammatory disturbances in CD patients, particularly in the active group. Hemoglobin, serum iron, albumin, and zinc were significantly lower in active disease, whereas folate and vitamin B12 did not differ significantly between groups. These findings are consistent with prior reports describing frequent anemia, iron deficiency, hypoalbuminemia, and zinc deficiency in patients with Crohn’s disease, especially during active inflammation [10,23,24]. However, biochemical markers in CD require cautious interpretation, as albumin is a negative acute-phase reactant, and hemoglobin, iron, and zinc may be influenced by inflammation, blood loss, malabsorption, disease location, treatment exposure, and supplementation. The weak-to-moderate overall correlations between BMI and these markers, together with the loss of most correlations after subgroup stratification, reinforce the notion that anthropometry complements biochemical analysis as a supportive but not interchangeable tool in nutritional assessment [25,26]. From a practical perspective, the present findings suggest that nutritional screening, preferably by a multimodal approach, should not be confined to hospitalized or overtly symptomatic patients but should also form part of routine outpatient follow-up, including remission visits, to enable earlier identification and correction of nutritional deficits.

This study has several strengths. Disease activity was classified using complementary clinical, endoscopic, and imaging criteria rather than symptoms alone, and nutritional status was evaluated using a multidimensional approach that combined GLIM-based classification, anthropometric measurements, questionnaire-based screening, and biochemical assessments. The study also provides contemporary data from a South Asian tertiary-care setting where comparable evidence remains limited. However, several limitations should be acknowledged. The cross-sectional design precludes causal inference, the single-center tertiary-care setting may limit external generalizability, and advanced body-composition techniques were not available to directly quantify muscle mass. In addition, the MNA-SF is not a CD-specific instrument, and some biochemical measures may have been influenced by recent treatment exposure, supplementation, or the inflammatory response itself. Future multicenter longitudinal studies should incorporate direct body composition assessment, standardized dietary evaluation, and outcome-based follow-up to determine how nutritional impairment evolves over time and whether targeted nutritional intervention improves the disease course. In current clinical practice, routine nutritional screening at diagnosis and during follow-up, combined with targeted correction of micronutrient deficiencies and early dietetic support, appears justified. A structured nutritional assessment strategy should therefore be integrated into the routine management of Crohn’s disease in both active and remission phases.

## Data Availability

The dataset generated and/or analyzed during the current study is not publicly available because it contains potentially sensitive information, and public sharing could compromise participant confidentiality. De-identified data relevant to the findings of this study may be made available to qualified researchers upon reasonable request and subject to approval by the Institutional Review Board of Bangladesh Medical University, Shahbagh, Dhaka. Requests for data access should be directed to the Institutional Review Board at, referencing study registration number 4764 (approved on 11 January 2024). Data will be shared in accordance with participant consent, institutional policies, and applicable ethical and legal requirements.

## ACKNOWLEDGMENTS

The authors would like to express their gratitude to the Department of Biochemistry, Radiology, and Imaging at Bangladesh Medical University for their valuable assistance throughout this study. Special thanks are extended to Prof. Dr. Md. Razibul Alam, Department of Gastroenterology; Dr. Md. Shahed Morshed and Dr. Tania Tofail, Assistant Professor and PhD researcher in the Department of Endocrinology, Bangladesh Medical University, for their advice on statistical analysis and scientific editing.

## SUPPLEMENTARY MATERIAL

S1 Checklist. Completed STROBE Checklist

